# Longitudinal monitoring of respiratory syncytial virus, SARS-CoV-2 and influenza in wastewater of a Southern Indian city

**DOI:** 10.1101/2024.10.11.24315282

**Authors:** Apoorva Venkatesh, RS Sreelekshmi, Parishuddha Babu Movva, Manas K Madhukar, Aruna Panda, S. Venkata Mohan, Rakesh Mishra, Shivranjani C Moharir

## Abstract

The detection of diverse pathogens and chemical compounds in community wastewater facilitates the monitoring of public health trends of emerging diseases and health conditions. During the COVID-19 pandemic, detection of viral RNA in wastewater assisted in monitoring the infection rates in different geographies and this drew attention to the method of wastewater-based epidemiology (WBE). In contrast to individual clinical testing, WBE offers an affordable, population-wide overview of the infection status of a community including asymptomatic carriers and those without access to healthcare facilities. To understand the community status of the infections caused by respiratory syncytial virus, SARS-CoV-2 and influenza virus subtypes in the monsoon season in Vijayawada city in the state of Andhra Pradesh in India, we longitudinally analyzed wastewater samples once a week between July-August 2024 from 7 sewage treatment plants in the city. The data obtained from the multiplexed RT-qPCR was used to calculate the viral loads in the wastewater samples at the city level and the proportion of population shedding the virus was estimated. All three viruses RSV A+B, SARS-CoV-2 and Influenza A+B were detected in the wastewater during our sampling period. Amongst the three viruses, the city had the largest load of SARS-CoV-2 RNA in the wastewater followed by RSV A+B and Influenza A+B. The study demonstrates the potential of utilizing wastewater surveillance system coupled to multiplexed RT-PCR to understand the population level dynamics of co-existence of multiple pathogens during the monsoon season in the urban settings.

## 1 Introduction

Wastewater monitoring is a method traditionally employed to track the spread of pathogens, including viruses such as poliovirus and norovirus, in a community (Nelson et al., 1967; Sinclair et al., 2008). It has recently emerged as a valuable tool for community-wide surveillance of Corona Virus Disease 2019 (COVID-19) and antimicrobial resistance (Ahmed et al., 2020; Madhukar et al., 2024; Munk et al., 2022; Tharak et al., 2022). The COVID-19 pandemic has exposed numerous unforeseen realities. Within months of severe acute respiratory syndrome coronavirus-2 (SARS-CoV-2) infections spreading globally, most governments found themselves grappling with the fundamental task of determining the extent and geographical distribution of the infected populations. The presence of SARS-CoV-2 in the fecal matter has led several research groups worldwide to advocate for wastewater based epidemiology (WBE) as a means to evaluate the viral circulation among populations (Agrawal et al., 2021; Mallapaty, 2020). Studies have consistently revealed correlations between wastewater monitoring and clinical case data for various viruses including SARS-CoV-2, respiratory syncytial virus (RSV) and influenza, thus reinforcing the value of wastewater surveillance as an epidemiological instrument (Hughes et al., 2022; Wolfe et al., 2022). Numerous investigations have identified other respiratory and enteric viruses in wastewater samples, including enteroviruses A-D, rhinovirus, parainfluenza and metapneumovirus (Boehm et al., 2023; Brinkman et al., 2017). These detections have also aligned with clinical disease patterns, offering real-time information on the viral circulation.

The virtue of wastewater sample to serve as a composite sample for a large population, and to provide health status of the entire community gives it a unique advantage, particularly when individual testing may be restricted by logistical or resource limitations. The detection of viral RNA in wastewater can also function as an early warning sign of the infection surges in communities, often preceding clinical caseloads (Ahmed et al., 2021). The COVID-19 pandemic has highlighted the critical need for more accurate and versatile methods for estimating and understanding the spread of the virus at community level that can complement the public health surveillance systems. Since COVID-19 was a global health threat during the pandemic, the focus was on monitoring SARS-CoV-2, the causative virus. However, attention must now be shifted on including the surveillance of the traditional seasonal respiratory viruses that have reemerged after being temporarily suppressed by pandemic control measures and efforts should be directed towards understanding their patterns of emergence along with the cropping patterns of the other dormant viruses.

Influenza A and B viruses and RSV are particularly noteworthy, which account for the majority of respiratory infections along with SARS-CoV-2 (García-Arroyo et al., 2022). Influenza A and B viruses are primarily linked to seasonal influenza outbreaks, with different subtypes of influenza A causing worldwide influenza pandemics (Hutchinson, 2018). The persistence of the viral nucleic acids in wastewater and the associated patterns of viral shedding of influenza A and B suggest that wastewater surveillance can be a useful method for identifying and tracking seasonal flu outbreaks and pandemics (Minodier et al., 2015). Research indicates that people carrying these viruses can release around millions copies of the virus per gram of fecal matter (Arena et al., 2012). Given that, influenza viruses and RSV present symptoms similar to SARS-CoV-2, it is challenging to differentiate between these infections based on only the clinical presentations (Chung et al., 2021). Using molecular approaches, wastewater surveillance that involves the detection of viral RNA in sewage, can be utilized as a crucial tool for distinguishing between COVID-19 surges and seasonal outbreaks of other respiratory viruses at community level, allowing researchers and public health authorities to monitor multiple respiratory pathogens simultaneously. This could facilitate a judicial resource relocation to manage the infectious diseases effectively.

This study aims to evaluate, monitor, and compare the longitudinal RNA levels of the three respiratory viruses-influenza, respiratory syncytial virus, and SARS-CoV-2, simultaneously, in wastewater samples collected from seven sewage treatment plants (STPs) in the southern Indian city of Vijayawada, in the state of Andhra Pradesh. The study demonstrates the potential of using a multiplexed real time polymerase chain reaction (RT-PCR) based approach to simultaneously monitor the longitudinal patterns of these viruses and differentiate their burden at community level through wastewater-based epidemiology. It also demonstrates how wastewater surveillance of these viral pathogens can be used to get an estimate of the proportion of the population shedding the virus and can be utilized as a complementary approach to strengthen the public health surveillance system.

## 2 Methods

### 2.1 Sample collection and processing

The wastewater samples were collected once a week in between 20^th^ July 2024 to 24^th^ August 2024 from 7 sewage treatment plants (STPs) of Vijayawada city that has a population of around 1,467,000 as of September 2024 (Population, 2024). The city generates around 132 million liters per day (MLD) sewage and most of it is treated through the sewage treatments plants in the city (Corporation, 2024). The details of the sampling sites that receive majorly human influx, are given in Supplementary Table 1. Around 1 liter wastewater sample was collected in sterile bottle during the peak flow from each STP by grab sampling method, as per the procedures described earlier (Kopperi et al., 2021; Madhukar et al., 2024; Tharak et al., 2022). The samples were inactivated using sodium hypochlorite (20 ml of 0.1% for 1 liter sample) (Hemalatha et al., 2021). Following thorough mixing, 50 ml of the wastewater sample was aliquoted, and 4 g of polyethylene glycol 8000 (Sigma -Aldrich, Catalogue No. P5413) and 0.9 g of sodium chloride (SRL, Catalogue No. 33205) were added to the sample and then incubated at 4°C overnight. The samples were centrifuged for 30 minutes at 10,000 rpm at 4°C. The pellet was either utilized directly for viral RNA extraction with the QIAamp Viral RNA Mini Kit (Qiagen, Catalogue No. 52906) or stored at – 80° C till further processing. Samples handling and processing was carried out in a biosafety cabinet following appropriate biosafety measures.

### 2.2 Detection of respiratory virus by RT-PCR

The RNA extracted from the wastewater samples were used for multiplex RT-PCR for the detection of the respiratory viruses using the GenePath Dx RespiRFC + Influenza subtyping + PMMoV Detection and Quantitation RT-qPCR Reagents kit (Catalog number: GPDX-RFC-3T-PM-01-100). Each sample was run in triplicate and, a 3 tubes 12 plex (4 plex X 3 tubes) reaction was set up for every sample. Tube 1 contained RSV A+B (FAM channel), SARS-CoV2 (HEX/ VIC channel), Influenza A+B (ROX channel), and an internal control PMMoV (Cy5 channel). Tube 2 contained targets for Influenza A (ROX channel), Influenza A H1N1 pdm09 (FAM channel), Influenza A H3N2 (HEX/ VIC channel), and an internal control PMMoV (Cy5 channel). And tube 3 contained Influenza A H7 (FAM channel), Influenza A H5 (HEX/ VIC channel), Influenza B (ROX channel), and the internal control PMMoV (Cy5 channel) as targets. RT-PCR was set with control templates of known concentrations of all the targets for plotting the standard curves for the respective targets. 5ul of 0.5 copies/ ul, 5 copies/ ul, 50 copies/ ul, 500 copies/ ul and 5000 copies/ ul of the control templates were used in triplicate for setting up RT-PCR reaction for obtaining the standard curves. The Ct values were plotted on Y axes against viral copies/ ul on the X axes. The equations obtained from the standard curves of each of the targets were used to calculate the respective viral RNA copies in the wastewater samples.

### 2.3 Statistical analysis

Based on the known concentrations of the controls and their respective Ct values, standard curves were plotted for the RNA concentrations for each of the viruses. An equation in the form of ‘y = mx + c’, where y = Ct value, m= gradient or slope, x = viral copies and c = y intercept, was obtained for each of the viruses. Pearson’s correlation co-efficient (R^2^) was calculated between the trend line and the line obtained by joining the datapoints. A correlation coefficient of ≥ 0.9 indicates a very strong positive correlation between the trend line and the line obtained by joining the datapoints.

### 2.4 Calculations of the estimated number of individuals shedding the virus

The total viral RNA load in the STP was estimated by multiplying the viral RNA copies per liter of wastewater by the total flow (MLD: Million Liters per Day) of wastewater processed in the treatment plant on that particular day (Supplementary Table 2 and 3).

*Total viral RNA load in the STP = viral RNA load per liter of wastewater * the volume of the STP*

Once the total viral RNA load in the STP is known then the estimated number of individuals contributing to this viral load can be calculated if the viral copies shed by an individual in feces is known (Ahmed et al., 2020; Hemalatha et al., 2021).

Based on literature for SARS-CoV-2, a single infected individual sheds 10^7^ copies of the viral RNA per ml of feces. Volume of feces excreted per day is 120 ml (Foladori et al., 2020; Hemalatha et al., 2021).

For influenza, literature reports that an infected individual sheds on an average 844700 viral RNA copies per gram of stool (Supplementary Table 4). This value is obtained by averaging the viral copies shed per gram of feces across different samples (Arena et al., 2012). On an average one person is known to excrete 128 grams or of feces per day (Rose et al., 2015)

The number of infected individuals contributing to the viral load in the STP is estimated by dividing the total viral load in the STP by the viral load shed by one infected individual per day:

*Number of individuals shedding the virus = Total viral load in STP/ (viral copies per gram or liter stool * gram or liter of stool excreted per day)*

## 3 Results

### 3.1 Simultaneous detection of RNA of respiratory viruses RSV A + B, SARS-CoV-2 and influenza in wastewater samples

Seven STPs in Vijayawada city were monitored once a week for the respiratory viruses-RSV A + B, SARS-CoV-2 and Influenza A + B between 20^th^ July 2024 to 24^th^ August 2024. Out of the 42 wastewater samples analyzed, 26 had Ct values ≤ 35 for RSV A + B, 41 had Ct ≤ 35 for SARS-CoV-2 and 32 had Ct ≤ 35 for Influenza A + B. PMMoV was used as an internal control and was detected in all the samples at Ct ≤ 35. 24 samples had Ct ≤ 35 for all the three viruses-RSV A + B, SARS-CoV-2 and Influenza A + B together (Table 1).

**Table 1:** Table showing the Ct values obtained by RT-PCR of wastewater samples for RSV A+B, SARS-CoV-2, influenza A+B, influenza A, influenza A H1N1, influenza A H3N2, influenza A H5, influenza A H7, influenza B and PMMoV. The Ct values in red font are ≤ 35. UD= Undetermined

| Collection date | Sample ID | RSV A+B | SARS-CoV2 | INF A+B | INF A | INF A H1N1 | INF A H3N2 | INF A H5 | INF A H7 | INF B | PMMoV |
| --- | --- | --- | --- | --- | --- | --- | --- | --- | --- | --- | --- |
| 20/07/24 | ASN1 | 36.70 | 32.21 | 35.23 | 38.57 | 38.18 | UD | UD | UD | UD | 26.15 |
|  | ASN2 | 37.02 | 32.69 | 36.70 | 37.20 | 37.74 | UD | UD | UD | UD | 26.92 |
|  | ATN1 | 35.21 | 31.51 | 33.86 | 36.70 | 37.16 | UD | UD | UD | UD | 25.80 |
|  | ATN2 | UD | 31.48 | 32.35 | 34.35 | 38.14 | UD | UD | UD | 33.77 | 25.16 |
|  | RLN1 | 35.10 | 31.55 | 34.03 | 36.58 | 38.05 | UD | UD | UD | 34.91 | 25.60 |
|  | RLN2 | 32.76 | 30.71 | 33.20 | 35.66 | 36.14 | UD | UD | UD | 34.97 | 25.12 |
|  | JKP | UD | 30.37 | 30.96 | 37.04 | 37.53 | UD | UD | UD | 37.02 | 25.31 |
| 27/07/24 | ASN1 | 34.60 | 28.44 | 34.19 | 37.07 | 36.91 | UD | UD | UD | 36.43 | 24.93 |
|  | ASN2 | 35.63 | 27.77 | 33.49 | 35.55 | 37.14 | UD | UD | UD | 36.21 | 24.24 |
|  | ATN1 | 36.30 | 29.29 | 36.31 | 38.91 | 37.76 | UD | UD | UD | 35.41 | 27.74 |
|  | ATN2 | UD | 32.16 | 35.84 | 38.39 | 38.18 | UD | UD | UD | UD | 28.00 |
|  | RLN1 | 36.25 | 29.62 | 34.28 | 36.04 | 38.21 | UD | UD | UD | 36.10 | 26.87 |
|  | RLN2 | 36.32 | 30.90 | 34.46 | 37.42 | 37.73 | UD | UD | UD | 37.28 | 27.62 |
|  | JKP | 37.64 | 28.23 | 36.66 | 37.19 | 37.71 | UD | UD | UD | 36.14 | 25.52 |
| 03/08/24 | ASN1 | 32.45 | 30.47 | 33.42 | 34.99 | 35.66 | UD | UD | UD | 34.08 | 25.13 |
|  | ASN2 | 32.41 | 29.78 | 32.71 | 34.01 | 36.36 | UD | UD | UD | 32.95 | 25.24 |
|  | ATN1 | 33.77 | 30.29 | 33.28 | 33.91 | 34.35 | UD | UD | UD | 33.57 | 24.83 |
|  | ATN2 | 34.15 | 31.12 | 35.37 | 35.76 | UD | UD | UD | UD | 34.65 | 26.22 |
|  | RLN1 | 34.74 | 31.39 | 34.28 | 35.62 | 37.51 | UD | UD | UD | 34.24 | 27.00 |
|  | RLN2 | 34.70 | 30.27 | 33.58 | 34.99 | 37.98 | UD | UD | UD | 34.61 | 25.75 |
|  | JKP | 32.54 | 30.21 | 33.10 | 36.32 | 37.69 | UD | UD | UD | 33.16 | 24.73 |
| 10/08/24 | ASN1 | 32.97 | 32.23 | 34.68 | 36.38 | 37.03 | UD | UD | UD | 35.36 | 25.44 |
|  | ASN2 | 35.41 | 33.17 | 35.62 | 36.44 | 38.53 | UD | UD | UD | 36.09 | 27.34 |
|  | ATN1 | 36.09 | 31.61 | 34.38 | 36.10 | 37.49 | UD | UD | UD | 34.88 | 24.42 |
|  | ATN2 | 33.50 | 29.64 | 33.86 | 35.15 | 37.58 | UD | UD | UD | 35.77 | 24.63 |
|  | RLN1 | 34.23 | 31.00 | 33.78 | 35.27 | 38.65 | UD | UD | UD | 34.60 | 25.03 |
|  | RLN2 | 34.92 | 32.98 | 35.13 | 37.43 | 38.78 | UD | UD | UD | 34.77 | 27.26 |
|  | JKP | UD | 36.59 | UD | 40.00 | UD | UD | UD | UD | UD | 32.82 |
| 17/08/24 | ASN1 | 34.65 | 30.57 | 33.76 | 33.92 | 34.60 | UD | UD | UD | 34.16 | 24.49 |
|  | ASN2 | 33.47 | 30.17 | 33.36 | 33.13 | 34.60 | UD | UD | UD | 33.63 | 24.33 |
|  | ATN1 | 32.69 | 30.64 | 34.30 | 35.19 | 36.83 | UD | UD | UD | 34.02 | 24.58 |
|  | ATN2 | 34.42 | 30.38 | 34.88 | 33.77 | 36.43 | UD | UD | UD | 34.07 | 23.54 |
|  | RLN1 | 35.14 | 30.55 | 35.51 | 35.31 | 35.16 | UD | UD | UD | 34.95 | 23.96 |
|  | RLN2 | 34.70 | 29.92 | 33.96 | 34.40 | 35.84 | UD | UD | UD | 33.29 | 24.82 |
|  | JKP | 34.71 | 30.32 | 34.62 | 35.74 | 36.30 | UD | UD | UD | 33.61 | 24.61 |
| 24/08/24 | ASN1 | 34.27 | 31.96 | 33.32 | 34.15 | 37.05 | UD | UD | UD | 31.40 | 24.41 |
|  | **ASN2** | 34.88 | 32.80 | 34.62 | 35.86 | 36.90 | UD | UD | UD | 33.28 | 26.21 |
|  | **ATN1** | 34.78 | 32.52 | 34.76 | 35.47 | 36.13 | UD | UD | UD | 34.29 | 25.89 |
|  | **ATN2** | 33.39 | 31.29 | 34.13 | 34.64 | 36.62 | UD | UD | UD | 33.43 | 24.83 |
|  | **RLN1** | 34.34 | 31.43 | 34.29 | 35.53 | 37.04 | UD | UD | UD | 34.28 | 25.73 |
|  | **RLN2** | 34.91 | 32.59 | 34.21 | 35.88 | 35.74 | UD | UD | UD | 33.55 | 26.39 |
|  | **JKP** | 34.51 | 31.01 | 32.84 | 33.06 | 34.13 | UD | UD | UD | 33.19 | 24.53 |

### 3.2 Longitudinal monitoring of RSV A+B, SARS-CoV-2 and Influenza A+B viral RNA in wastewater

RSV A+B, SARS-CoV-2 and Influenza A+B viral RNA were detected by RT-PCR in most of the samples analyzed during the monsoon period of 20^th^ July 2024 to 24^th^ August 2024. In order to calculate the viral RNA concentrations in the wastewater samples from the Ct values, we plotted the standard curves for each of the viruses using control templates of known concentrations (Figure 2 A-J). Using the Ct values obtained for each of the known concentrations, an equation was derived for the respective viruses. We then calculated the concentration of the viral RNA in the respective STPs on the dates of sample collection using the Ct values obtained from the RT-PCR of the RNA extracted from the wastewater samples and fitting them in the standard curve equations for the respective virus. To understand the longitudinal patterns of these viruses, the viral RNA concentrations were plotted against the date of sampling (Figure 3-5). Amongst the three respiratory viruses-RSV A+B, SARS-CoV-2 and Influenza A+B, the RNA load was maximum for SARS-CoV-2 in general and on the peak dates as compared to the other two.

**Figure 1:**
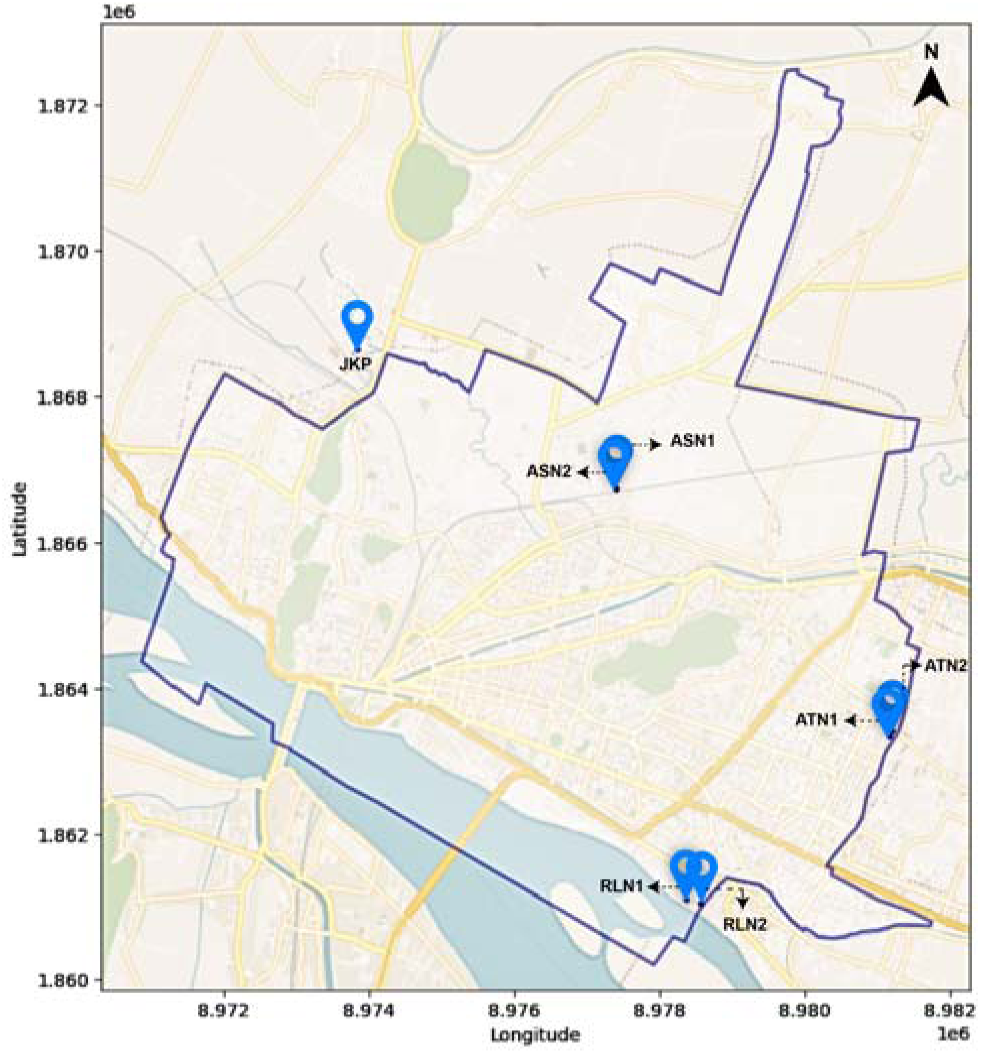
Co-ordinates of the sampling sewage treatment plants plotted on the map of Vijayawada city.

**Figure 2:**
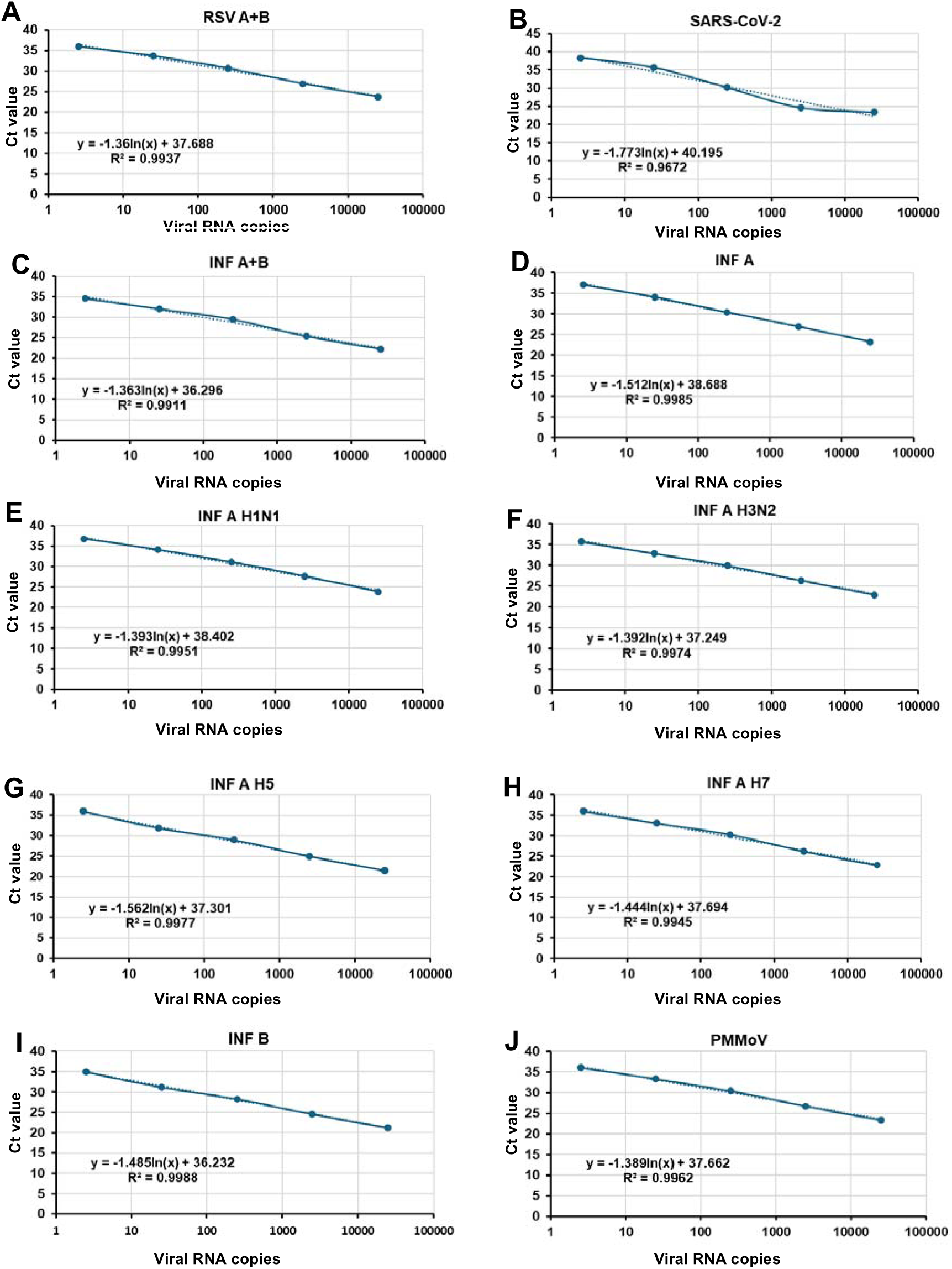
Standard curves of viruses, -A) RSV A+B, B) SARS CoV-2, C) INF A+B, D) INF A, E) INF A H1N1, F) INF A H3N2, G) INF A H5, H) INF A H7, I) INF B, and J) PMMoV.

**Figure 3:**
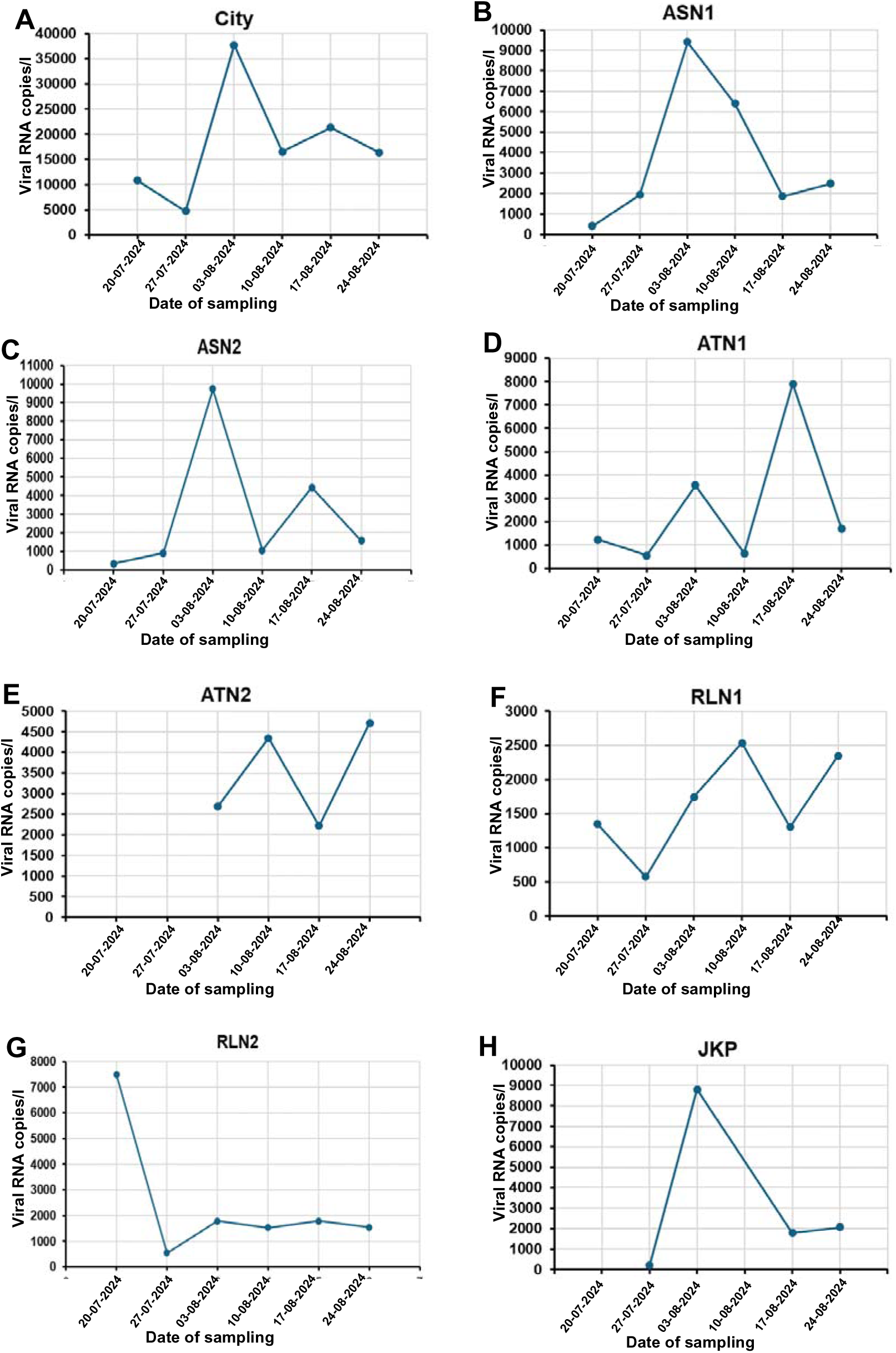
Longitudinal plots of RNA load of RSV in wastewater samples A) At the city level, and the 7 sampling locations, - B) at the location ASN1 -, C) at the location ASN2, D) at the location ATN1, E) at the location ATN2, F) at the location RLN1, G) at the location RLN2 and H) at the location JKP.

For RSV A+B, there was a clear rise in the viral RNA load in the city in the third sampling week, i.e., on 3^rd^ August 2024 (Figure 3A). This increase in RSV A+B RNA load on 3^rd^ August was also observed in the individual sampling locations ASN1, ASN2 and JKP, while for the location RLN1, the RNA load peaked on 10^th^ August 2024 (Figure 3 B, C, F and H). The location ATN1 showed the rise in RSV A+B RNA load on 17^th^ August 2024, which declined in the subsequent weeks (Figure 3D). The RSV A+B RNA was not detected in the location ATN2 on 20^th^ July and 27^th^ July 2024 and the location JKP on 20^th^ July 2024 (Figure 3E and H). In the location RLN2, the RSV A+B RNA load was higher in the first week of sampling, i.e., on 20^th^ July 2024 and declined in the subsequent weeks (Figure 3G).

SARS-CoV-2 RNA load peaked in the second week of sampling i.e., on 27^th^ July 2024 during the sampling window in the city (Figure 4A). The peak of SARS-CoV-2 RNA load was also seen in the individual locations ASN1, ASN2, ATN1 RLN1 and JKP on 27^th^ July 2024 (Figure 4 B, C, D, F and H). In the location ATN2, the peak was seen on 10^th^ August 2024, while in RLN2 it was seen on 17^th^ August 2024 (Figure 4E and G).

**Figure 4:**
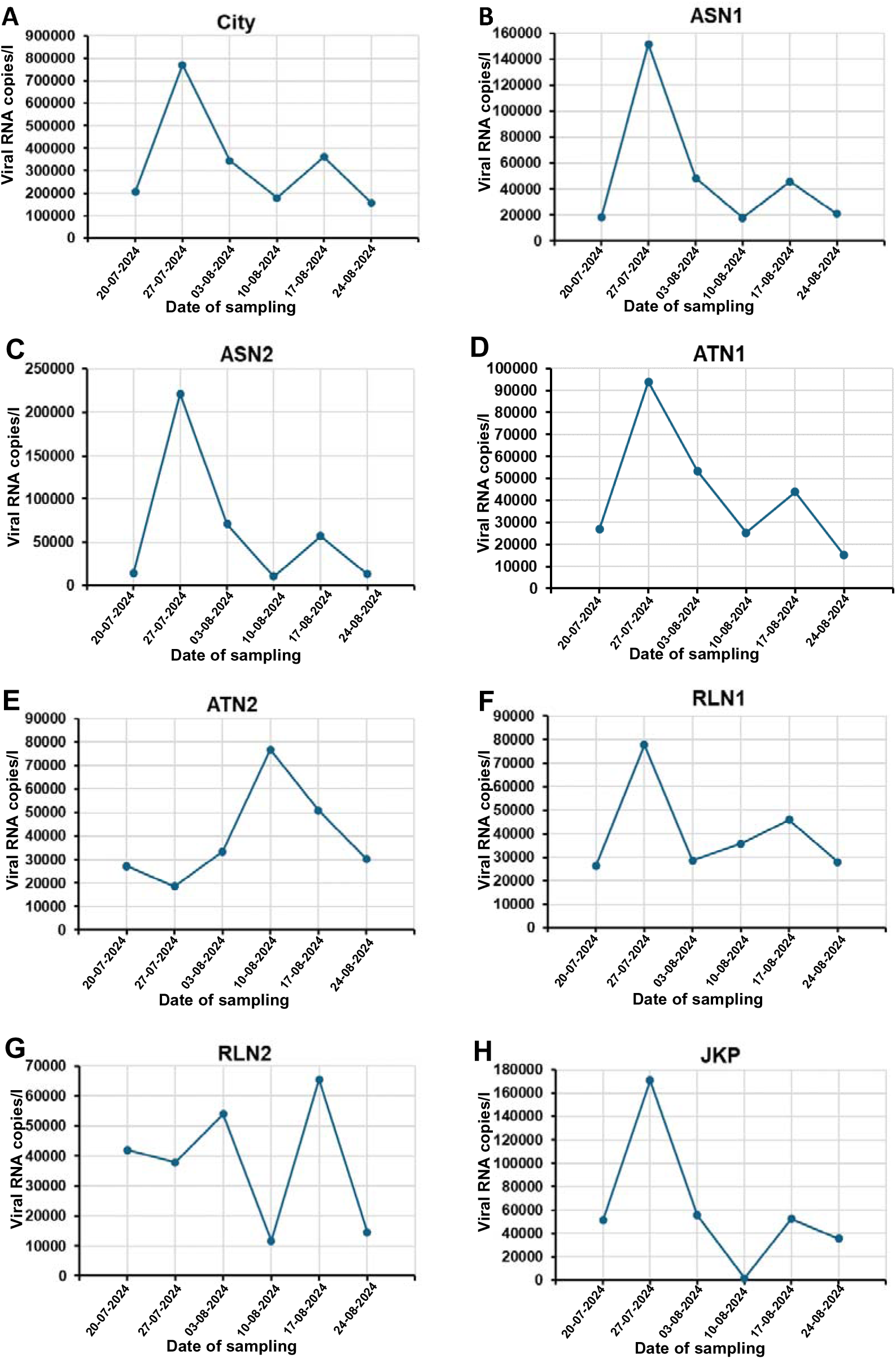
Longitudinal plots of RNA load of SARS-CoV-2 in wastewater samples A) At the city level, and the 7 sampling locations, B) at the location ASN1 -, C) at the location ASN2, D) at the location ATN1, E) at the location ATN2, F) at the location RLN1, G) at the location RLN2 and H) at the location JKP.

The RNA load of Influenza A+B virus was highest in the city on 20^th^ July 2024 and had maximum contribution from the site JKP, ATN2 and RLN2 on that day (Figure 5A, E, G and H). The locations ASN2 and ATN1 showed the peek on 3^rd^ August 2024 (Figure 5C and D). The location ASN1 showed two peaks-one on 3^rd^ August and the other on 24^th^ August 2024 during the sampling window, while the location RLN1 showed the peak on 10^th^ August 2024 (Figure 5B and F).

**Figure 5:**
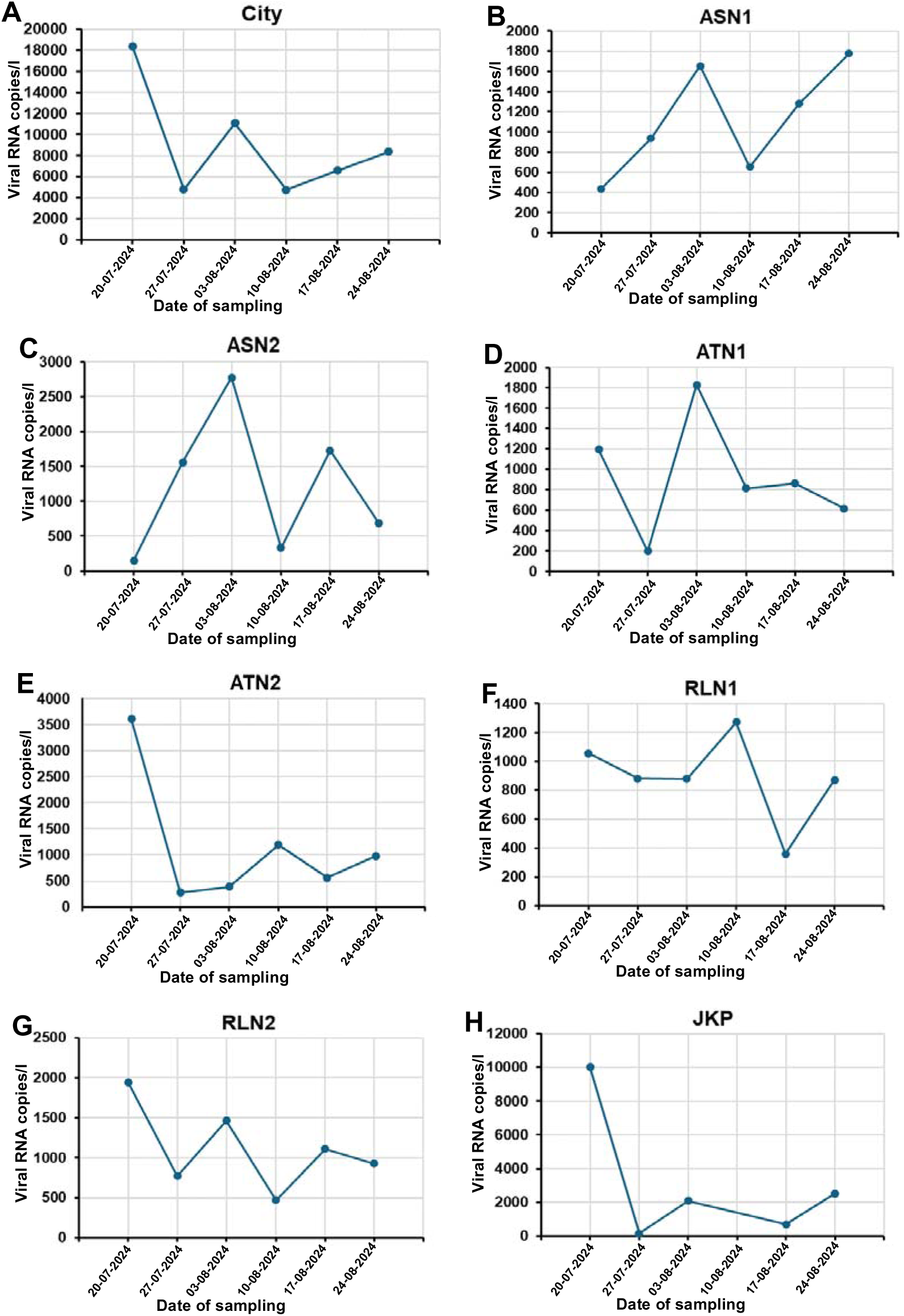
Longitudinal plots of RNA load of Influenza in wastewater samples A) At the city level, and the 7 sampling locations, B) at the location ASN1 -, C) at the location ASN2, D) at the location ATN1, E) at the location ATN2, F) at the location RLN1, G) at the location RLN2 and H) at the location JKP.

### 3.3 Detection of subtypes of influenza through wastewater surveillance

Since the Influenza viral RNA was detected in all the locations in the city during the sampling period, we investigated the presence of the RNA of different subtypes of Influenza virus viz Influenza A virus and its subtypes -Influenza A H1N1, Influenza A H3N2, Influenza A H5 and Influenza A H7, as well as Influenza B virus. 12 out of 32 Influenza A + B samples with ≤ 35 had Ct value ≤ 35 for primer-probes specific to Influenza A as well, of which 4 samples showed Ct ≤ 35 for Influenza A H1N1. Influenza A H3N2, Influenza A H5 and Influenza A H7 were not picked up in any of the samples. 27 samples showed Ct value ≤ 35 for Influenza B (Table 1). The longitudinal trend plots for the RNA load of Influenza A, Influenza A H1N1 and Influenza B are shown in supplementary figures 1-3.

### 3.4 Estimation of number of infected individuals

The viral RNA copies excreted in feces by an individual infected with SARS-CoV-2 and Influenza virus is known in literature. Combining this information with the viral load that observed in the individual STPs in our study, we estimated the number of individuals shedding the viral RNA into the wastewater in the respective individual STPs and the whole city (Table 2 and 3). We observed that during our sampling window, on 20^th^ July 2024, 0.21% population of the city was shedding the SARS-CoV-2 RNA in the wastewater (Figure 6A). The percentage of individuals shedding the SARS-CoV-2 RNA in wastewater increased to 0.94% in the next week, followed by a decline in subsequent weeks. A more granular analysis at individual location level suggested that the location ASN1 had the maximum SARS-CoV-2 RNA shedding in wastewater, by 0.344 % of the population, on 27^th^ July 2024, followed by 0.251% at the location ASN2 and 0.194% at the location JKP on the same day (Figure 6C).

**Figure 6:**
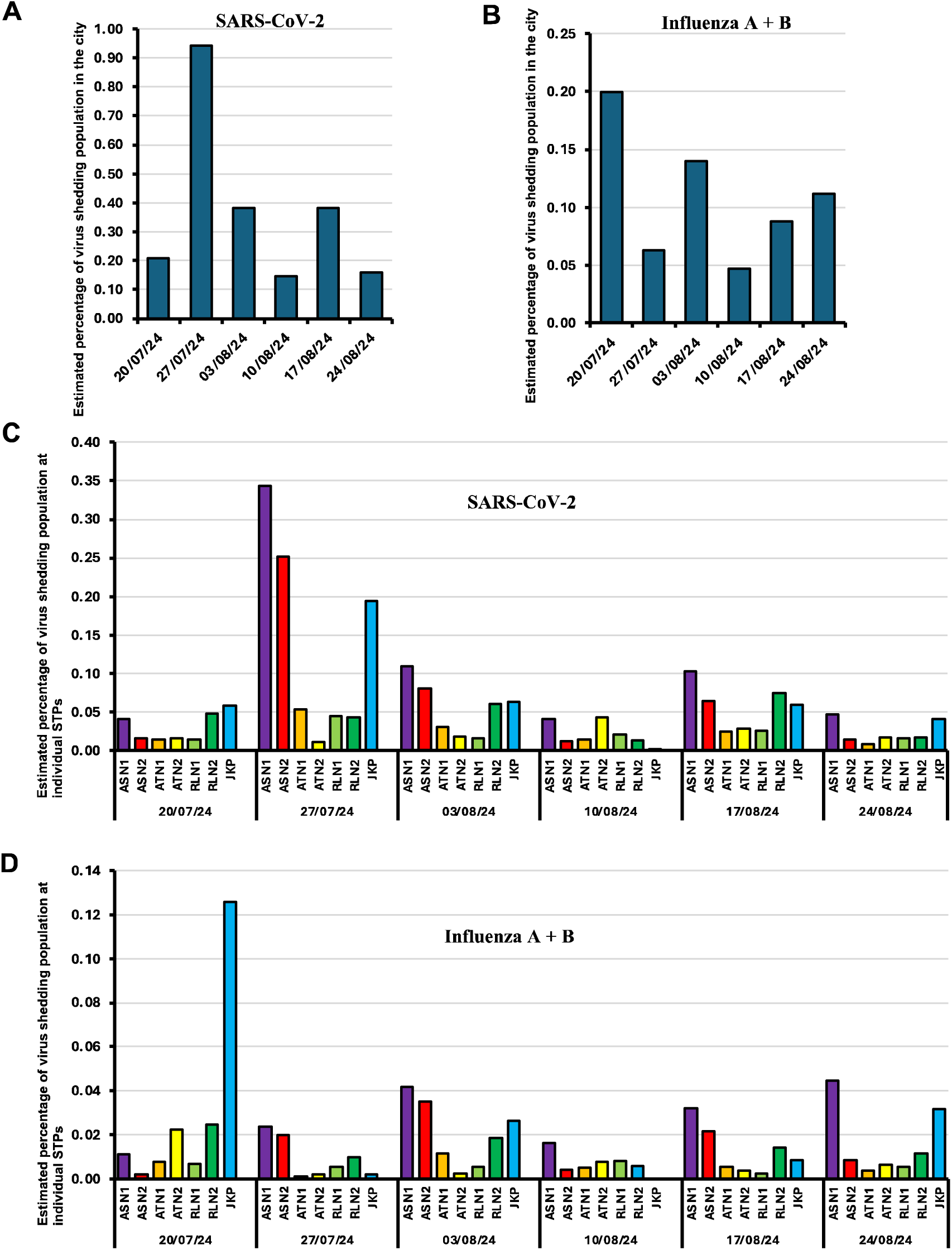
Estimated percentage of virus shedding population A) Estimated percentage of SARS-CoV-2 shedding population at the city level, B) Estimated percentage of Influenza A+B shedding population at the city level, C) Estimated percentage of SARS-CoV-2 shedding population at the 7 individual STPs and A) Estimated percentage of Influenza A+B shedding population at the 7 individual STPs.

**Table 2:** Table showing the SARS-CoV-2 RNA load in wastewater samples, the estimated number of infected individuals and the estimated percentage of infected individuals in the city.

| Collection date | STP | SARS-CoV-2 RNA in wastewater sample (copies/l) | Estimated number of infected individuals | Estimated percentage of infected individuals in the city |
| --- | --- | --- | --- | --- |
| 20/07/24 | ASN1 | 18098.16 | 603 | 0.041 |
|  | ASN2 | 13753.78 | 229 | 0.016 |
|  | ATN1 | 26855.92 | 224 | 0.015 |
|  | ATN2 | 27242.88 | 227 | 0.015 |
|  | RLN1 | 26234.19 | 219 | 0.015 |
|  | RLN2 | 41990.41 | 700 | 0.048 |
|  | JKP | 51060.08 | 851 | 0.058 |
|  | **Total** | **205235.42** | **3053** | **0.208** |
| 27/07/24 | ASN1 | 151472.89 | 5049 | 0.344 |
|  | ASN2 | 221066.78 | 3684 | 0.251 |
|  | ATN1 | 93899.71 | 782 | 0.053 |
|  | ATN2 | 18574.85 | 155 | 0.011 |
|  | RLN1 | 77723.00 | 648 | 0.044 |
|  | RLN2 | 37824.57 | 630 | 0.043 |
|  | JKP | 170908.98 | 2848 | 0.194 |
|  | **Total** | **771470.78** | **13797** | **0.941** |
| 03/08/24 | ASN1 | 48269.27 | 1609 | 0.110 |
|  | ASN2 | 71260.30 | 1188 | 0.081 |
|  | ATN1 | 53458.68 | 445 | 0.030 |
|  | ATN2 | 33431.84 | 279 | 0.019 |
|  | RLN1 | 28652.69 | 239 | 0.016 |
|  | RLN2 | 53942.23 | 899 | 0.061 |
|  | JKP | 55647.88 | 927 | 0.063 |
|  | **Total** | **344662.90** | **5586** | **0.381** |
| 10/08/24 | ASN1 | 17824.67 | 594 | 0.041 |
|  | ASN2 | 10481.96 | 175 | 0.012 |
|  | ATN1 | 25400.46 | 212 | 0.014 |
|  | ATN2 | 76792.06 | 640 | 0.044 |
|  | RLN1 | 35788.12 | 298 | 0.020 |
|  | RLN2 | 11690.55 | 195 | 0.013 |
|  | JKP | 1523.79 | 25 | 0.002 |
|  | **Total** | **179501.61** | **2139** | **0.146** |
| 17/08/24 | ASN1 | 45651.60 | 1522 | 0.104 |
|  | ASN2 | 57042.80 | 951 | 0.065 |
|  | **ATN1** | 43884.52 | 366 | 0.025 |
|  | **ATN2** | 50829.28 | 424 | 0.029 |
|  | **RLN1** | 45962.46 | 383 | 0.026 |
|  | **RLN2** | 65526.86 | 1092 | 0.074 |
|  | **JKP** | 52457.21 | 874 | 0.060 |
|  | **Total** | **361354.71** | **5611** | **0.382** |
| **24/08/24** | **ASN1** | 20783.28 | 693 | 0.047 |
|  | **ASN2** | 12968.10 | 216 | 0.015 |
|  | **ATN1** | 15202.12 | 127 | 0.009 |
|  | **ATN2** | 30315.02 | 253 | 0.017 |
|  | **RLN1** | 28042.03 | 234 | 0.016 |
|  | **RLN2** | 14568.96 | 243 | 0.017 |
|  | **JKP** | 35551.75 | 593 | 0.040 |
|  | **Total** | **157431.26** | **2357** | **0.161** |

**Table 3:** Table showing the Influenza RNA load in wastewater samples, the estimated number of infected individuals and the estimated percentage of infected individuals in the city.

| Sample collection date | STP | Influenza RNA in wastewater sample (copies/l) | Estimated number of infected individuals | Estimated percentage of infected individuals in the city |
| --- | --- | --- | --- | --- |
| 20/07/24 | ASN1 | 435.94 | 161 | 0.011 |
|  | ASN2 | 148.32 | 27 | 0.002 |
|  | ATN1 | 1196.58 | 110 | 0.008 |
|  | ATN2 | 3603.65 | 332 | 0.023 |
|  | RLN1 | 1055.97 | 97 | 0.007 |
|  | RLN2 | 1944.42 | 358 | 0.024 |
|  | JKP | 10017.57 | 1846 | 0.126 |
|  | **Total** | **18402.45** | **2933** | **0.200** |
| 27/07/24 | ASN1 | 937.01 | 345 | 0.024 |
|  | ASN2 | 1563.21 | 288 | 0.020 |
|  | ATN1 | 198.09 | 18 | 0.001 |
|  | ATN2 | 279.28 | 26 | 0.002 |
|  | RLN1 | 880.60 | 81 | 0.006 |
|  | RLN2 | 770.90 | 142 | 0.010 |
|  | JKP | 153.60 | 28 | 0.002 |
|  | **Total** | **4782.69** | **929** | **0.063** |
| 03/08/24 | ASN1 | 1650.42 | 608 | 0.041 |
|  | ASN2 | 2778.36 | 512 | 0.035 |
|  | ATN1 | 1830.73 | 169 | 0.012 |
|  | ATN2 | 395.30 | 36 | 0.002 |
|  | RLN1 | 878.19 | 81 | 0.006 |
|  | RLN2 | 1463.59 | 270 | 0.018 |
|  | JKP | 2083.85 | 384 | 0.026 |
|  | **Total** | **11080.44** | **2060** | **0.140** |
| 10/08/24 | ASN1 | 653.33 | 241 | 0.016 |
|  | ASN2 | 328.74 | 61 | 0.004 |
|  | ATN1 | 815.76 | 75 | 0.005 |
|  | ATN2 | 1194.42 | 110 | 0.008 |
|  | RLN1 | 1270.80 | 117 | 0.008 |
|  | RLN2 | 469.92 | 87 | 0.006 |
|  | JKP |  | 0 | 0.000 |
|  | **Total** | **4732.97** | **690** | **0.047** |
| 17/08/24 | ASN1 | 1282.85 | 473 | 0.032 |
|  | ASN2 | 1726.08 | 318 | 0.022 |
|  | ATN1 | 862.89 | 80 | 0.005 |
|  | **ATN2** | 564.21 | 52 | 0.004 |
|  | **RLN1** | 356.91 | 33 | 0.002 |
|  | **RLN2** | 1110.39 | 205 | 0.014 |
|  | **JKP** | 686.01 | 126 | 0.009 |
|  | **Total** | **6589.35** | **1287** | **0.088** |
| **24/08/24** | **ASN1** | 1775.40 | 654 | 0.045 |
|  | **ASN2** | 686.34 | 127 | 0.009 |
|  | **ATN1** | 618.34 | 57 | 0.004 |
|  | **ATN2** | 981.26 | 90 | 0.006 |
|  | **RLN1** | 872.69 | 80 | 0.005 |
|  | **RLN2** | 926.46 | 171 | 0.012 |
|  | **JKP** | 2523.79 | 465 | 0.032 |
|  | **Total** | **8384.27** | **1645** | **0.112** |

During our sampling window, we observed the maximum RNA shedding of Influenza virus in the wastewater in the city, corresponding to shedding by 0.2% of the population, on 20^th^ July 2024 (Figure 6 B). The location JKP had the maximum percentage of individuals, 0.126%, contributing to the shedding of viral RNA on that day (Figure 6D). There was a decrease in the shedding of viral RNA in the wastewater in the subsequent weeks.

Except for the location JKP on 2^th^ July 2024, at any point of time, the percentage of population shedding SARS-CoV-2 in wastewater was higher as compared to those shedding Influenza A + B at city level as well as individual location level. The concentration of the RSV RNA shed by an infected individual in the feces is not known. Hence, we could not estimate the number of individuals shedding the virus in the STPs wastewater.

## Discussion

Though the prominent symptoms shown by the individuals infected with the respiratory viruses RSV, SARS-CoV-2 and Influenza RNA are related to the respiratory systems, the RNA of these viruses are shed in the wastewater through the fecal route as well. Earlier studies have demonstrated that there is a strong correlation between the shedding of viral RNA by infected individuals in wastewater and the respective number of clinically confirmed individuals (Tisza et al., 2023; Toribio-Avedillo et al., 2023). In this study, we investigated the levels of these three viruses together in wastewater in Vijayawada. All the three viruses could be detected in the wastewater in the city. The load of SARS-CoV-2 RNA was the highest in the city throughout our sampling window, followed by RSV A+ B and the Influenza A + B.

The influenza virus consists of 4 distinct types of viruses-influenza A, B, C and D. While influenza A primarily affects the humans as well as other hosts including aquatic birds, pigs, horses and poultry, influenza B and C mainly infects humans, and influenza D is known to infect cattle, pigs and goats (Francis, 1940; Toribio-Avedillo et al., 2023). It is well understood that influenza A and B viruses co-occur in humans across different age groups and exhibit varying degrees of severity (He et al., 2022). Annually, influenza A and B viruses contribute to between 291,243 and 645,832 respiratory-related deaths, with 3 to 5 million cases of severe illness occurring worldwide (Iuliano et al., 2018).

In India, a nation of 1.428 billion people, the surveillance capacity for influenza A H1N1 virus is limited, with only few laboratories equipped to conduct clinical testing according to National Centre for Disease Control (NCDC, 2019). In such scenario, wastewater and sewage surveillance can offer considerable potential for the detection of influenza viruses and their subtypes. A recent pan-India study that investigated 34,260 cases of influenza-like illness (ILI) and severe acute respiratory infection found that during the 2021 monsoon season in India, influenza A(H3) and B/Victoria strains predominated. However, in the 2022 monsoon season, influenza A (H1N1) pdm09 became the dominant strain (Potdar et al., 2023). Concordantly, in our study, we could detect the H1N1 subtype in wastewater in the monsoon season of July and August 2024. The avian influenza A subtypes H3N2, H5, and H7 were not detected in any of the samples during the sampling window. This could be possible due to four reasons-i) in this study, we sampled the sewage treatment plants that receive influx from human catchments and not from poultry, so these subtypes if present, could not find a way in the sampled wastewater, ii) due to absence of these infectious during our sampling window, iii) inefficient or improbable detection of these subtypes through wastewater surveillance or iv) probably these subtypes are not shed in the feces. H5N1 caused significant outbreaks of avian flu in Asian countries in 2004, H7N3 in Pakistan, and H5N2 in Taiwan, leading to the culling of hundreds of millions of poultry (Sturm-Ramirez et al., 2004). Further investigations are necessary towards detection of these subtypes in wastewater surveillance and to optimize the protocols.

Interestingly, in our wastewater samples, influenza B virus was detected in 27 of the 42 samples (Ct value ≤ 35), representing a relatively higher proportion compared to influenza A. Unlike influenza A, which can infect both humans and animals, influenza B is a strictly human pathogen, having evolved stably to infect humans. This likely explains its higher detection rate in sewage samples (Chen and Holmes, 2008).

The presence of SARS-CoV-2 in wastewater has been documented in numerous studies and has garnered significant attention within the WBE community (Zhang et al., 2022). While much attention has been focused on WBE for tracking SARS-CoV-2, this approach has also demonstrated its effectiveness in detecting and monitoring a variety of enteric viruses like influenza, RSV, poliovirus, norovirus, rotaviruses, human astroviruses, and hepatitis A and E viruses. To date, there has been minimal research on the concurrent detection of RSV, Influenza, and SARS-CoV-2 in wastewater in India. Our study represents the comprehensive investigation in India to simultaneously monitor the presence of SARS-CoV-2, RSV, and influenza viruses in wastewater and provides valuable insights into the co-circulation of these pathogens within the community.

## Data Availability

All data produced in the present work are contained in the manuscript

---

**Graphical abstract.**
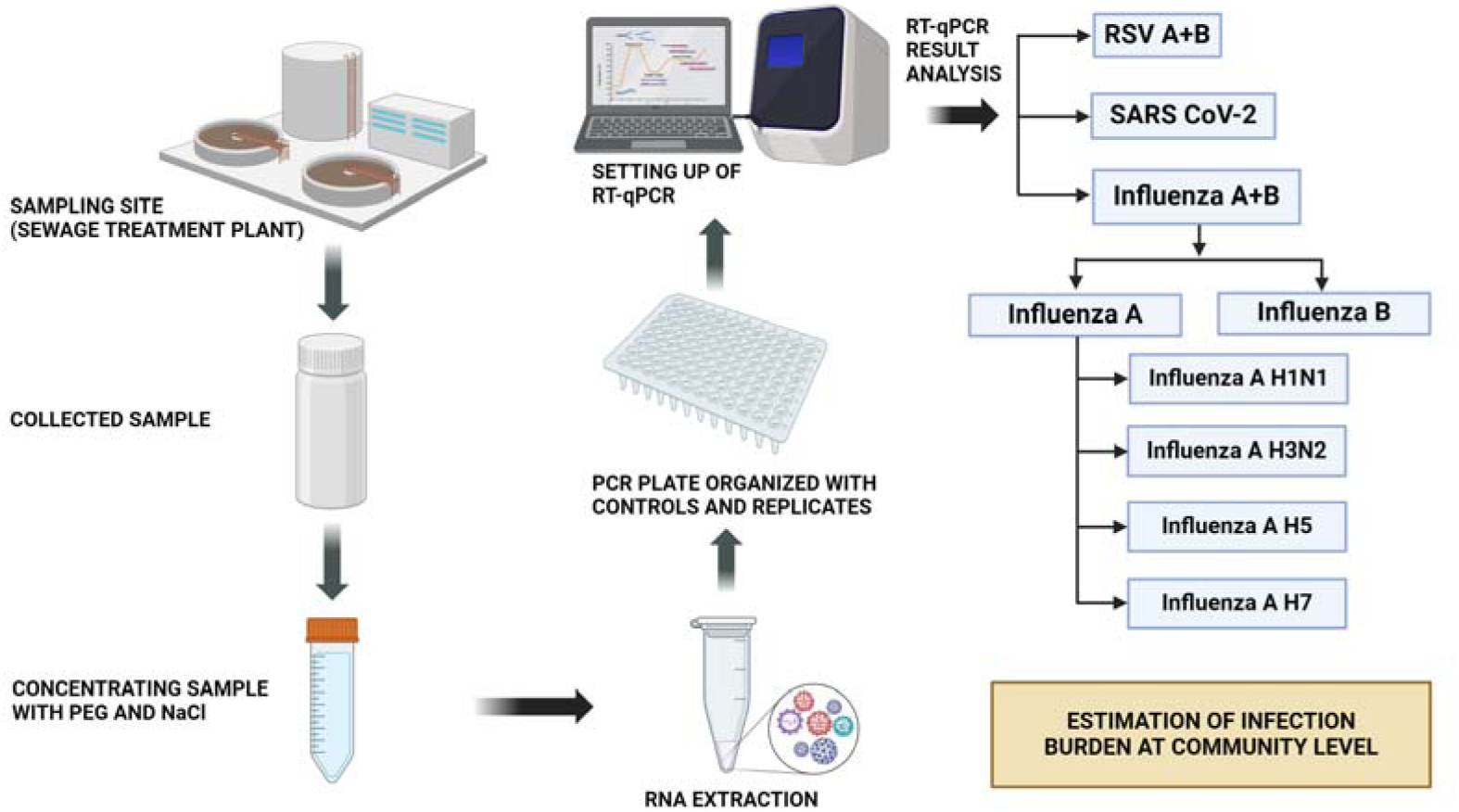

## Abbreviations

COVID-19: Corona Virus Disease 2019
SARS-CoV-2: sever acute respiratory syndrome coronavirus -2
WBE: wastewater based epidemiology
RSV: respiratory syncytial virus
STPs: sewage treatment plants
RT-PCR: real time polymerase chain reaction
MLD: Million Liters per Day

## Author contributions

SCM, RKM, VM and AP conceptualized and planned the study. RKM and SCM received the funding for the study. AV, SRS, PBM, MKM performed the experiments. SCM, AV, SRS, MKM performed the data analysis and wrote the first draft of the manuscript. All the authors contributed to the refinement of the manuscript draft.

## Acknowledgements

The authors acknowledge the generous intellectual support of Dr. L S Shashidhara, Director, NCBS, Bengaluru and appreciate his inputs on the study.

